# Diarrheal disease attributed to *Shigella* spp. and Enteroinvasive *Escherichia coli* among children at households in rural Haïti: A case-control study

**DOI:** 10.1101/2025.01.29.25321342

**Authors:** Ishae Sriguha, Saradiya R Kuyt, Youseline Cajusma, Emilee Cato, Lindsey Brinkley, Md Abu Sayeed, Stace Maples, Valery Madsen Beau De Rochars, Daniel T Leung, Anthony T Maurelli, Chantale Baril, Molly B Klarman, Eric J Nelson

**Author notes:** Co-first authors (Ishae Sriguha, Saradiya R Kuyt). **Corresponding authors** Ishae Sriguha, Eric Nelson.

## Abstract

Infections from *Shigella* spp. and Enteroinvasive *Escherichia coli* (EIEC) are considered leading causes of symptomatic diarrheal disease, globally. However, there is a paucity of case-control studies from Caribbean nations to guide regional public health priorities and interventions. A case-control study was conducted within a larger cross-sectional healthcare study in Haïti. Participant households were identified using a geospatially randomized method; families with children under 5 years were consented and enrolled. Rectal swabs from child participants were tested for *Shigella* spp./EIEC by qPCR using the *ipaH* target. Two case-definitions were used: ‘diarrheal symptom’ (DS) cases were defined as those reporting diarrheal symptoms ≤7 days ago; ‘acute diarrhea’ (AD) cases were defined as those reporting diarrheal symptoms ≤7 days ago with ≥3 loose stools in the past 24 hours and onset <7 days ago. Of 868 households screened, 568 were enrolled with 794 participating children; samples from 732 children were analyzed. Rates of *Shigella* spp./EIEC. detection among DS cases and controls were 11.4% (22/193) and 6.1% (33/539), respectively. Rates of detection among AD cases and controls were 18.6% (8/43) and 6.8% (47/689), respectively. The adjusted odds of having DS increased by 84% (aOR=1.84; 95%CI 1.02 to 3.27) and having AD increased by 183% (aOR=2.83; 95%CI 1.14 to 6.36) when *Shigella* spp./EIEC was detected. The attributable fractions for DS and AD were 5.62% (95%CI 0.44% to 10.9%) and 12.6% (95%CI 0% to 25.3%), respectively. Rates of bloody diarrhea (dysentery) were minimal (<1%, 6/732). Within this case-control study in Haïti, *Shigella* spp./EIEC detection was common and attributed to symptomatic disease. These results align with prior global health studies. *Shigella* spp./EIEC represent an important public health target for intervention once the security situation in Haïti stabilizes.

**AUTHOR SUMMARY:** Global studies outside the Caribbean have identified a group of closely related enteric pathogens (*Shigella* spp., Enteroinvasive *Escherichia coli* (EIEC)) as a leading cause of diarrheal disease. This group can be associated with short-term (e.g., bloody diarrhea) or long-term (e.g., malnutrition, stunting) health impacts. Within the Caribbean, there is a lack of information to guide public health policy on enteric pathogens, especially in Haïti. Therefore, we conducted a case-control study where we identified households with children under 5 years, gathered data on diarrhea symptoms, and collected stool swabs. We tested the stools using a sensitive molecular assay (*ipaH* qPCR). *Shigella* spp./EIEC detection was common among both children with and without diarrhea, yet more common among, and attributed to, those with symptomatic diarrheal disease. These results align with prior global health studies and represent an opportunity for high-impact public health action when the security situation in Haïti stabilizes.

## INTRODUCTION

Diarrheal diseases are the second leading cause of death among children between one month and 5 years of age worldwide with at least 1.1 billion cases and 446,000 deaths annually (1). Morbidity has both acute components and more complex bi-directional correlates that include malnutrition, stunting, and neuro-cognitive delay (2–5). *Shigella* spp./EIEC are a group of closely related bacterial pathogens estimated to cause 111 million cases and 63,100 deaths annually among children under 5 years (5–8).

The pathophysiologic consequences of infection vary. Upon fecal-oral transmission, *Shigella* spp. infection may manifest in symptomatic diarrheal disease with or without blood in the stool (9). Fluid loss leading to dehydration occurs at the level of the small intestine (9). Invasive disease leading to enterocolitis generally occurs at the level of the large intestine in a subset of patients (10). While shigellosis can be self-limited, antibiotics are indicated for severe disease and vulnerable sub-populations (11). The infectious dose can be as low as 10-100 bacilli which likely contributes to large outbreaks when hygiene and sanitation infrastructure are compromised (7, 12). The Pan American Health Organization (PAHO) considers *Shigella* spp. to be one of the most dangerous antibiotic resistant pathogens in Latin America and the Caribbean, exhibiting increased rates of resistance to antibiotics such as ciprofloxacin and azithromycin (4, 13, 14).

A landmark multisite case-control study (‘GEMS’) found that *Shigella* spp./EIEC were the leading enteric pathogens detected. The GEMS study found the attributable fraction for *Shigella* spp./EIEC to range from 3.9% to 83.2% depending on age stratum and study site (15). *Shigella* spp./EIEC was the most attributable or one of the most attributable pathogens in infants, toddlers, and older children (15). A second landmark multisite birth cohort study (‘MAL-ED’), among children ≤ 2 years with and without diarrheal disease who were not seeking care, found an attributable fraction of 10.2% for *Shigella* spp./EIEC (16). While both were global in reach, there was only a single site in Latin America (Peru), and there remains a paucity of insight from case-control studies from the Caribbean, including Haïti, to set regional public health priorities.

The taxonomy of this group of pathogens is complicated because approaches were established in the pre-genomics era. Phylogenetics revealed *Shigella* spp. and EIEC are closely related (17). *Shigella* spp. are Gram-negative, nonmotile, facultatively anaerobic, non-spore-forming, rod-shaped bacteria (18). They are differentiated from *Escherichia coli* by pathophysiology and metabolism (e.g., inability to ferment lactose) (18). The Shigella genus is divided into four serogroups with multiple serotypes: A (*S. dysenteriae*, 12 serotypes); B (*S. flexneri*, 6 serotypes); C (*S. boydii*, 18 serotypes); and D (*S. sonnei*, 1 serotype); these pathogens can readily be detected by PCR/qPCR using primers for the invasion plasmid antigen H gene (*ipaH*) (9, 19). Given that *ipaH* is also found among Enteroinvasive *Escherichia coli* (EIEC) it is difficult to disaggregate these pathogens without cost-prohibitive molecular and culture analyses (19). Both the GEMS and MAL-ED studies chose to treat *ipaH* positive samples as a singleton entity when assessing attributions to symptomatic infection.

We hypothesized that among children in Haïti, *Shigella* spp./EIEC would be detected at rates similar to the GEMS and MAL-ED studies and the odds of *Shigella* spp./EIEC detection would be higher among, and attributed more often to, those with diarrheal disease. To test these hypotheses, we conducted a case-control study among children at the household level who were enrolled within a larger cross-sectional study in Haïti, a setting with progressively deteriorating public health infrastructure, security and governance (20). Despite limited data at the household level, diarrheal diseases have been studied among care seeking children in Haïti (21–23). In 2012, *Shigella* spp. were detected in 21.5% and 18.2% of children under 5 years presenting at a cholera treatment center and an oral rehydration point in Port au Prince, respectively (23). Our findings were consistent with our guiding hypothesis and aligned well with both the GEMS and MAL-ED studies (15, 16).

## METHODS

### Ethics statement

Ethical approvals were obtained from the Institutional Review Board at the University of Florida (IRB201703246) and the Comité National de Bioéthique (National Bioethics Committee of Haïti, Ref:1718-35).

### Study population and setting

A case-control study was conducted within a larger cross-sectional study investigating healthcare-seeking intention and behavior in Haïti (20). The study population resides within a deteriorating healthcare infrastructure due to socio-political unrest (24). The under-five mortality rate is approximately 56 deaths per 1000 live births and the stunting rate is 20% (25). In 2023, rates of vaccination of 12 vaccines in the WHO/UNICEF schedule ranged from only 41-75%, with some vaccine coverage lower than in 2013, possibly due to the state of unrest (26). Participant households were located in the communes of Gressier and Leogane Haïti (located 12 and 27 miles respectively southwest of the capital Port-au-Prince). The enrollment strategy aimed to equitably enroll families across rural, peri-urban and urban areas. The study area (477 km^2^) was divided into grid cells two square kilometers in size and each cell was classified by density; “low” (0-112 structures), “medium” (113-397 structures), or “high” (398-4664 structures). Six grid cells of each density were randomly selected for sampling. Each of the selected grid cells was divided into 18, 24, or 53 Thiessen polygons based on their designation as low, medium, or high density, respectively. The study team navigated to a targeted polygon and screened the first house they encountered. This process was repeated until at least one house in each polygon was enrolled or all households were screened. Enrollment criteria included a head of household who consented to complete a household survey and had at least one child under five years old for whom consent was obtained to collect a stool swab. The surveys were conducted to ascertain the socioeconomic status of families and the recent diarrhea history of child participants. Although not part of the criteria, participants were not actively seeking care at the time of enrollment.

### Clinical procedures

Anthropomorphic measurements were obtained including height, weight, and mean upper arm circumference (MUAC). Temperature was measured and dehydration status was assessed using WHO IMCI methods for children reporting current fever or diarrhea symptoms, respectively (27).

### Laboratory procedures

#### Sample collection

Stool samples were obtained via rectal swabs, immediately placed on ice in transport tubes with 3 mL of normal saline and brought to a laboratory in Gressier. The contents of the transport tubes were divided into two 2 mL tubes and stored at −80°C. Samples were transported on dry ice to the Emerging Pathogens Institute at University of Florida for analysis.

#### Molecular assays

Standard methods for nucleic acid extraction were performed (28, 29). NEB Luna Universal qPCR Master Mix, nuclease free water, and primers (*ipaH* or *16S*) were combined as a master mix. The *ipaH* primer was chosen for the detection of *Shigella* spp./EIEC and the *16S* primer was used as a *eubacteria* control (30, 31). Each well of a Hard-Shell® 96-Well PCR plate was loaded with 20 µL of master mix along with either 1 µL of sample or 1 µL of control. The plates were set onto a thermocycler (Bio Rad CFX96 Touch Real-Time PCR Detection System) to denature at 95°C for one minute, then 35 cycles of denaturing at 95°C for 15 seconds, and annealing and extension at 60°C for 30 seconds. Two operators conducted two biological replicates for each of nine 96-well plates and switched between two thermocycler machines (except for plate 2) to control for machine or operator bias. One positive control of *Shigella* spp. genomic DNA and two negative controls of nuclease-free water were included on each plate.

### Analytic strategy

#### Definitions

Two definitions for cases and controls were established based on survey responses to questions about history of diarrheal symptoms (Fig S1). Cases for ‘diarrheal symptoms’ (DS) were defined as answering ‘YES’ to the question “Diarrheal symptoms within the last 7 days?”. Controls for ‘diarrheal symptoms’ were defined as answering ‘NO’ to the same question. Cases for ‘acute diarrhea’ (AD) were defined as answering ‘YES’ to the previous question as well as the questions “Three or more loose stools in the past 24 hours?” and “Onset less than 7 days ago?”. Controls for ‘acute diarrhea’ were defined as answering ‘NO’ to any or all of the three previous questions.

#### Analysis of molecular data

Results from the qPCR reactions were analyzed quantitatively and qualitatively. Blank readings indicative of no detection of *ipaH* were set to the maximum possible Ct value of 35. Qualitatively, samples were ruled positive if the *ipaH* Ct was less than 28 and the *16S* Ct value was less than 26. The *ipaH* threshold was based on standard curves and positive/negative controls; the *16S* threshold was based on distributions among experimental samples and positive/negative controls. Samples negative for *16S* were excluded from further analysis.

#### Statistical analysis

A socio-economic status (SES) index was previously generated using components from the household survey (20). Data were described by proportions for categorical variables and medians with interquartile ranges for continuous variables. Characteristics of cases were compared to controls within each case definition using the two-sided non-paired Mann-Whitney U test, Chi-square test for independence, and Fisher’s exact test. For the primary analysis, an odds ratio was used to compare odds of *Shigella* spp./EIEC detection among cases and controls for both DS and AD case definitions. Additionally, adjusted odds ratios were calculated to account for age, sex, and socioeconomic status. Attributable fractions were calculated with the graphPAF package in R using the Bruzzi formula with 1,000 bootstrap replications (32, 33). A two-sided non-paired Mann-Whitney U test was used to compare differences in Ct values between cases and controls for each definition for all samples and only *ipaH* positive samples. All analyses were performed using R Statistical Software (v4.4.1) (34, 35). Figures were created using R Statistical Software (v4.4.1) and GraphPad Prism (v10.4.1) (34, 36).

## RESULTS

### Participant characteristics

Of the 868 households screened, 568 were enrolled with 794 children. Among the enrolled children, 732 met inclusion criteria for the case-control analysis (Fig 1). Over half of the participants were between 24-59 months (57%, n=416) and the median age was 26 months (IQR 12-42 months) (Tables 1 and 2). Participation by sex was approximately equal (51%, n=372 female). The median MUAC was 146 mm (IQR 139-155 mm) and based on these measurements 1% (n=5) were severely malnourished (Fig 2) (37). The majority of both male and female participants were within 3 z-scores of their age-specific median weight (98%, 708; Fig S2) and median height (93%, 681; Fig S3) (38). Approximately 10% (n=71) of participants reported a current fever, 1% (n=6) reported blood in stool and none of the participants with acute diarrhea (0/43) were dehydrated (Table 3).

**Fig 1.**
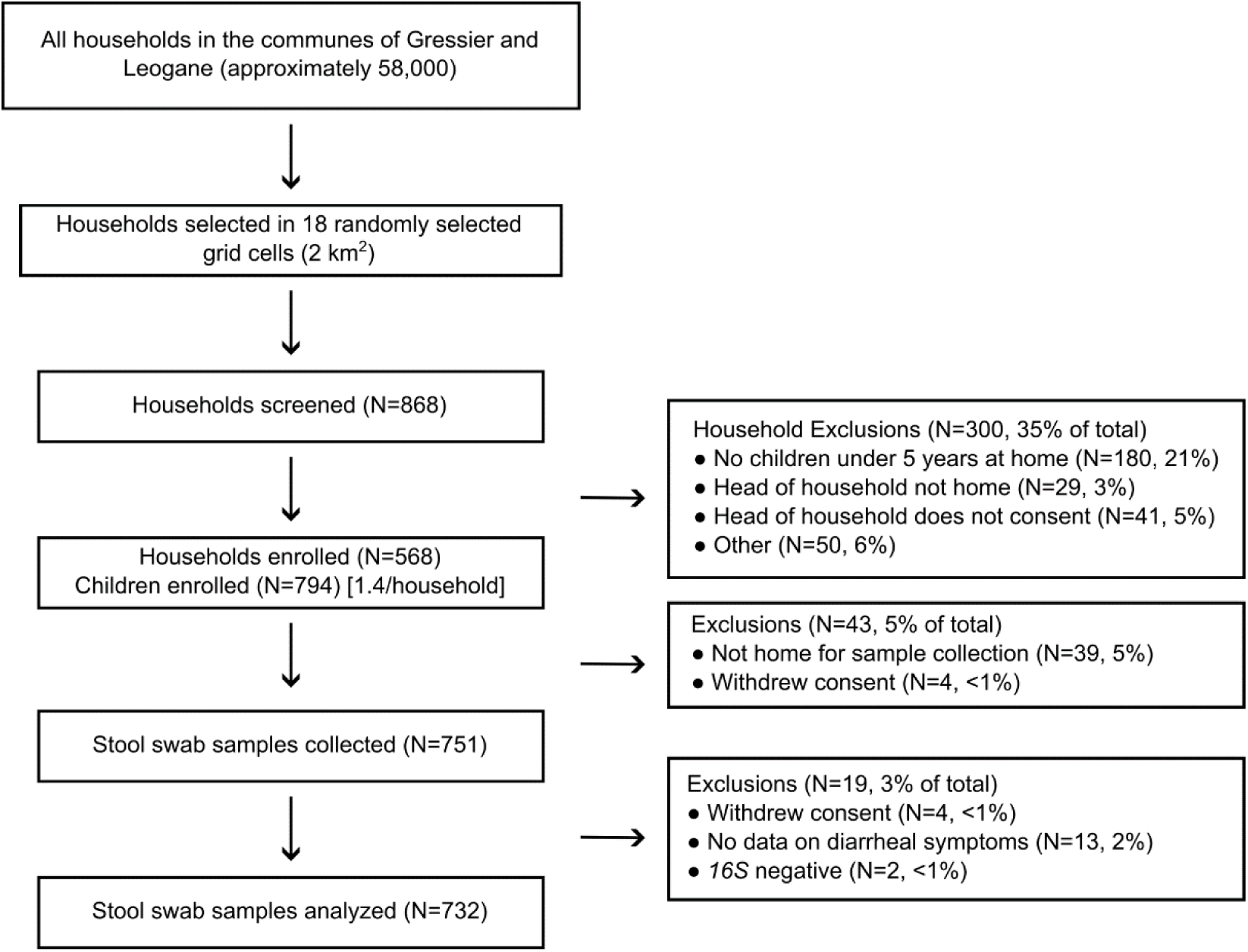
Participant enrollment and exclusions.

**Fig 2.**
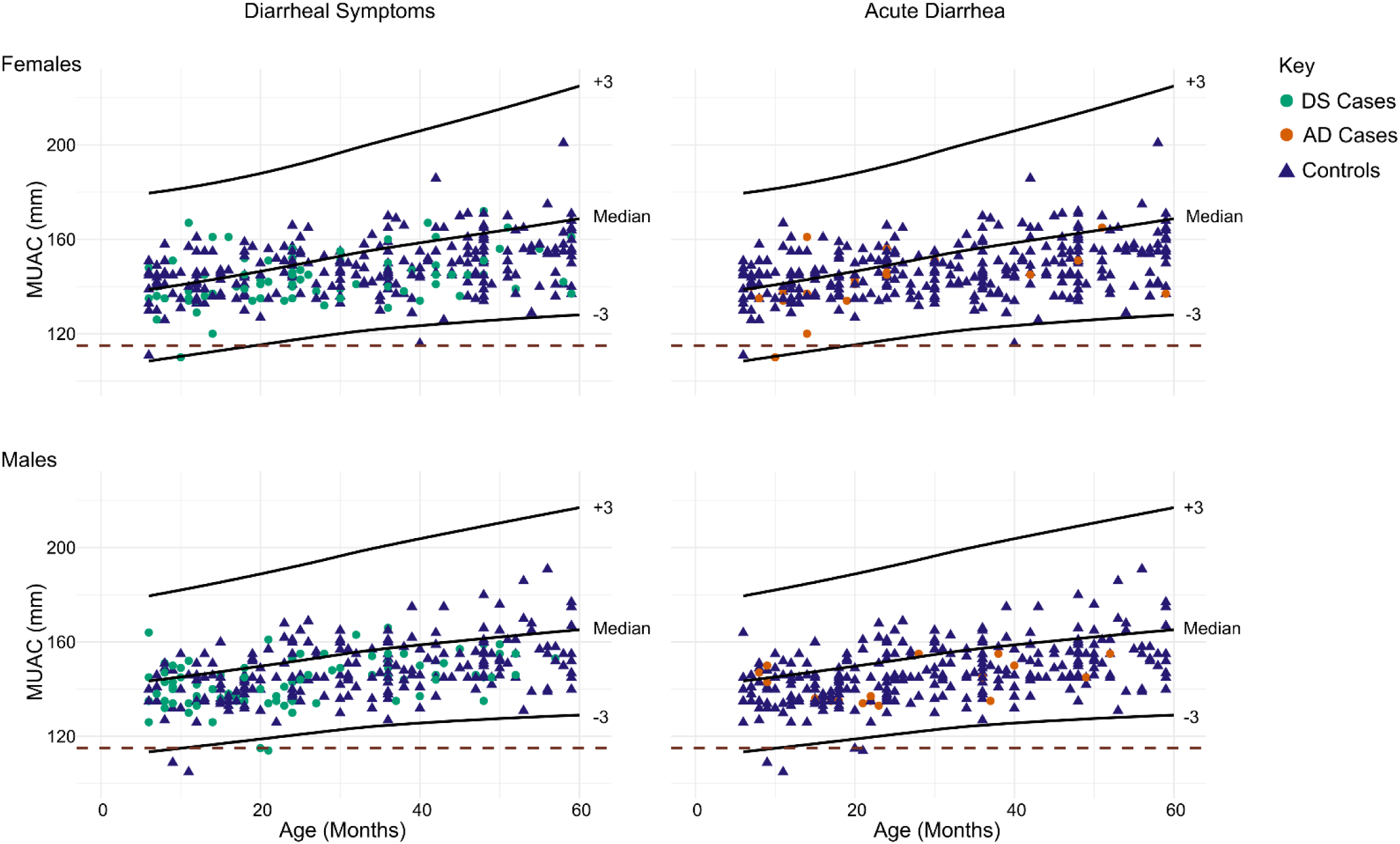
Sex-specific MUAC measurements and Z-scores for all participants as well as stratifications for DS and AD case definitions. A MUAC <115 mm is considered severely malnourished. DS=Diarrheal symptoms; answered ‘YES’ to “diarrheal symptoms within the last 7 days”. AD=Acute diarrhea; answered ‘YES’ “diarrheal symptoms within the last 7 days”, and “three or more loose stools in the past 24 hours”, and “onset less than 7 days ago”. MUAC=mid-upper arm circumference.

**Table 1.** Identification of factors associated with diarrhea symptoms (DS)

|  | Cases | Controls | OR<sup>a</sup> (95% CI) | P | aOR<sup>b</sup> (95% CI) | P |
| --- | --- | --- | --- | --- | --- | --- |
| Diarrheal symptoms (DS)<sup>c</sup> | 193 (26%) | 539 (74%) |  |  |  |  |
| *ipaH* positive | 22 (11%) | 33 (6%) | **1.973 (1.106-3.457)** | **0.019** | **1.838 (1.017-3.265)** | **0.040** |
| Age; median months (IQR)<sup>d</sup> | 21 (10–36) | 30 (14.5-46) | **0.976 (0.966-0.986)** | **<0.001** | - | - |
| Age |  |  |  |  |  |  |
| 0-11 months | 59 (31%) | 103 (19%) | REF | REF | REF | REF |
| 12-23 months | 49 (25%) | 105 (20%) | 0.815 (0.510-1.300) | 0.390 | 0.844 (0.523-1.358) | 0.485 |
| 24-59 months | 85 (44%) | 331 (61%) | **0.448 (0.301-0.669)** | **<0.001** | **0.456 (0.305-0.683)** | **<0.001** |
| Sex; Female | 90 (47%) | 282 (52%) | 0.796 (0.572-1.107) | 0.175 | 0.830 (0.591-1.163) | 0.278 |
| SES<sup>e</sup> Index |  |  |  |  |  |  |
| 1<sup>st</sup> quintile | 41 (21%) | 99 (18%) | REF | REF | REF | REF |
| 2<sup>nd</sup> quintile | 36 (19%) | 117 (22%) | 0.743 (0.440-1.251) | 0.264 | 0.793 (0.464-1.35) | 0.395 |
| 3<sup>rd</sup> quintile | 43 (22%) | 101 (19%) | 1.028 (0.617-1.714) | 0.915 | 1.120 (0.663-1.896) | 0.672 |
| 4<sup>th</sup> quintile | 40 (21%) | 108 (20%) | 0.894 (0.534-1.496) | 0.670 | 0.907 (0.535-1.535) | 0.715 |
| 5<sup>th</sup> quintile | 33 (17%) | 114 (21%) | 0.699 (0.409-1.187) | 0.187 | 0.680 (0.392-1.170) | 0.164 |
| MUAC (IQR) | 143 (135-150) | 146 (141-155) | **0.967 (0.952-0.981)** | **<0.001** | - | - |

11 <sup>a</sup> OR=odds ratio.12 <sup>b</sup> aOR= adjusted odds ratio. Age (Continuous) and MUAC were not accounted for when computing the adjusted odds ratios.13 <sup>c</sup> DS=Diarrheal symptoms; answered 'YES' to "diarrheal symptoms within the last 7 days".14 <sup>d</sup> IQR=Inter quartile range.15 <sup>e</sup> SES=Socioeconomic status. See methods for components of the index.

**Table 2.** Identification of factors associated with acute diarrhea (AD)

|  | Cases | Controls | OR<sup>a</sup> (95% CI) | P | aOR<sup>b</sup> (95% CI) | P |
| --- | --- | --- | --- | --- | --- | --- |
| Acute diarrhea (AD)<sup>c</sup> | 43 (6%) | 689 (94%) |  |  |  |  |
| *ipaH* positive | 8 (19%) | 47 (7%) | **3.122 (1.286-6.813)** | **0.007** | **2.832 (1.140-6.400)** | **0.016** |
| Age; median months (IQR)<sup>d</sup> | 20 (9.5-29) | 27 (13-43) | **0.977 (0.958-0.996)** | **0.019** | - | - |
| Age |  |  |  |  |  |  |
| 0-11 months | 14 (33%) | 148 (22%) | REF | REF | REF | REF |
| 12-23 months | 14 (33%) | 140 (20%) | 1.057 (0.483-2.314) | 0.888 | 1.050 (0.467-2.360) | 0.905 |
| 24-59 months | 15 (35%) | 401 (58%) | **0.395 (0.185-0.848)** | **0.016** | **0.412 (0.192-0.892)** | **0.023** |
| Sex; Female | 19 (44%) | 353 (51%) | 0.753 (0.401-1.397) | 0.371 | 0.826 (0.433-1.558) | 0.555 |
| SES<sup>e</sup> index |  |  |  |  |  |  |
| 1<sup>st</sup> quintile | 11 (26%) | 129 (19%) | REF | REF | REF | REF |
| 2<sup>nd</sup> quintile | 8 (19%) | 145 (21%) | 0.647 (0.244-1.648) | 0.365 | 0.744 (0.275-1.940) | 0.547 |
| 3<sup>rd</sup> quintile | 10 (23%) | 134 (19%) | 0.875 (0.353-2.144) | 0.769 | 1.040 (0.409-2.629) | 0.934 |
| 4<sup>th</sup> quintile | 4 (9%) | 144 (21%) | 0.326 (0.089-0.979) | 0.060 | 0.335 (0.090-1.025) | 0.070 |
| 5<sup>th</sup> quintile | 10 (23%) | 137 (20%) | 0.856 (0.346-2.097) | 0.732 | 0.841 (0.331-2.115) | 0.711 |
| MUAC | 142 (135-150) | 146 (140-155) | **0.967 (0.941-0.993)** | **0.015** | - | - |

<sup>a</sup> OR=odds ratio.<sup>b</sup> aOR= adjusted odds ratio.<sup>c</sup> AD=Acute diarrhea; answered 'YES' "Diarrheal symptoms within the last 7 days", and "Three or more loose stools in the past 24 hours", and "Onset less than 7 days ago".<sup>d</sup> IQR=Inter quartile range.<sup>e</sup> SES=Socioeconomic status. See methods for components of the index.
Age (Continuous) and MUAC were not accounted for when computing the adjusted odds ratios.

**Table 3.** Report of fever or blood in stool stratified by diarrheal symptom (DS) and acute diarrhea (AD) case definitions.

|  | All<br>n=732 | DS<sup>a</sup> Cases<br>n=193 | DS<sup>a</sup> Controls<br>n=539 | AD<sup>bc</sup> Cases<br>n=43 | AD<sup>bc</sup> Controls<br>n=689 |
| --- | --- | --- | --- | --- | --- |
| **Fever; n (%)** | 71 (10%) | 12 (6%) | 59 (11%) | 1 (2%) | 70 (10%) |
| **Blood in stool; n (%)** | 6 (1%) | 6 (3%) | 0 | 0 | 6 (1%) |

<sup>a</sup> DS=Diarrheal Symptoms; answered 'YES' to "diarrheal symptoms within the last 7 days".
<sup>b</sup> AD=Acute Diarrhea; answered 'YES' "diarrheal symptoms within the last 7 days", and "three or more loose stools in the past 24 hours", and "onset less than 7 days ago".
<sup>c</sup> Children with acute diarrhea were assessed for dehydration and all had 'no' dehydration.

### Characteristics of cases and controls

One quarter of participants (n=193) fit the case definition for ‘diarrheal symptoms’ (Table 1) and 6% (n=43) fit the more restrictive case definition for ‘acute diarrhea’ (Table 2). The DS and AD cases were slightly younger than the respective DS (P<.001) and AD (P=.022) controls and had slightly lower MUAC measurements than the respective DS (P=<.001) and AD (P=.010) controls. The proportion of cases who were female was similar to that of controls for both case definitions. There was no significant difference between cases and controls in the proportions of underweight or severely malnourished participants for either DS or AD case definitions. The SES distribution of DS and AD cases was not significantly different than their corresponding DS and AD controls.

### Pathogen detection

The overall rate of *ipaH* detection was 7.5% (n=55). Among the DS cases, 22 (11.4%) were *ipaH* positive, representing an increased odds of 97% (OR=1.97; 95%CI 1.11 to 3.46) for detection among participants with diarrheal disease (Table 1). Among the AD cases, 8 (5.9%) were *ipaH* positive, representing an increased odds of 212% (OR=3.12; 95% CI 1.29 to 6.81) for detection among participants with diarrheal disease (Table 2). When controlling for age, sex, and SES, detection of *ipaH* was associated with an increased odds of diarrheal disease for both case definitions; DS (aOR=1.84; 95% CI 1.02 to 3.27) and AD (aOR=2.83; 95% CI 1.14 to 6.36). Among participants with positive *ipaH* Ct values, the median value for DS cases (25.9; IQR 22.4-26.8) compared to DS controls (24.6; IQR 22.3-26.5) was not statistically different (p=0.23). The median *ipaH* Ct value among AD cases (26.4; IQR 24.3-26.9; p=.03) compared to AD controls (24.6; IQR 21.3-26.6) was slightly higher (p=0.03; Fig S4). The unadjusted attributable fractions of *Shigella* spp./EIEC for diarrheal disease were 5.62% (95% CI 0.44% to 10.9%) for DS and 12.6% (95% CI 0% to 25.3%) for AD. After adjusting for confounders, the attributable fractions of *Shigella* spp./EIEC to diarrheal disease were 5.2% (95% CI 0% to 10.5%) for DS and 12.0% (95% CI 0% to 24.8%) for AD.

## DISCUSSION

In this case-control study, detection of *Shigella* spp./EIEC was common and attributed to diarrhea symptoms among children under five years of age in the Caribbean nation of Haïti. Participants with *Shigella* spp./EIEC were twice as likely to report diarrheal symptoms (DS) within the seven-day period prior to rectal swab collection. Among participants meeting the more stringent acute diarrheal (AD) disease definition, the odds of having AD increased to three times as likely. When adjusting for the potential confounding variables of sex, age category, and SES, similar findings were made. Attributable fractions of *Shigella* spp./EIEC detection and symptomatic disease were more similar to the MAL-ED study than GEMS (15, 16). The GEMS study, stratified by age, found that *Shigella* spp./EIEC were the attributable pathogen in 3.9% to 15.8% of cases in infants, 15.4% to 64.9% of cases in toddlers, and 17.8% to 83.2% in older children across all sites (15). The difference in proportions compared to our findings may be due to differences in participant inclusion criteria, as those included in the GEMS study were actively seeking care (15). The MAL-ED study, while focusing on children under 2 years of age, included participants at the household level not actively seeking care. Our attributable fraction of 12.9% is close to their findings of 10.2% of cases that were attributed to *Shigella* spp./EIEC.

This study found negligible qPCR differences (<1 Ct) between cases and controls for both DS and AD definitions. This contrasts with the MAL-ED study that found participants with diarrhea attributed to *Shigella* spp./EIEC had lower Ct values (higher abundance) compared to controls as would be expected (15, 16, 39, 40). There are multiple explanations for our findings including study design (e.g., lower sample size), procedure (e.g. possible differences in extraction efficiencies) and biological variation (possible differences in target copy number between cases and controls). Importantly, our attributable fractions for *Shigella* spp./EIEC align well with the attributable fraction found in the MAL-ED study for diarrheal disease. Lastly, our study had low rates of reported blood in stool overall (< 1%). Whereas, in MAL-ED, 4.7% of participants with *Shigella*-specific diarrhea reported blood in stool (16). An underlying pathophysiologic explanation for this difference is unknown and would require additional laboratory studies to assess differences in underlying gut health between the two populations.

Our findings should be considered within the context of the study limitations. First, enrollment bias may have been present in the larger public health study (20) because recruitment occurred Monday to Friday during daytime hours when potentially eligible head of households and school aged children were likely outside of the home. Second, the case definitions for DS and AD relied on survey responses, which are not as accurate as data ascertained clinically. Third, there was a procedural delay of approximately five years between sample collection and analysis which poses a risk of sample degradation. To minimize this, *16S* primers were used as a control to confirm that expected rates of total bacteria were present in the stool sample and all *16S* negative samples were excluded. Further, the *ipaH* gene is present in both *Shigella* spp. and EIEC.(41) While EIEC infection is uncommon (42, 43), especially compared to other diarrheagenic pathogens including other *E. coli* pathogens and *Shigella* spp., they cannot be differentiated from *Shigella* spp. without further analysis (44). Serotyping can differentiate *Shigella* spp. from EIEC, however, antibodies to O antigens of *Shigella* spp. have cross-reactivity with EIEC O antigens (45, 46).

## CONCLUSION

Despite study limitations, *Shigella* spp./EIEC detection was common and associated with symptomatic disease among participants within the community. The results indicate the need for monitoring and control of *Shigella* spp./EIEC infections in Haïti as an etiological agent for diarrheal disease among children under five years. Intervention would reduce rates of acute disease as well as reduce nutritional and development sequalae associated with *Shigella* spp./EIEC.

## Acknowledgements

We thank the participants for joining in this study as well as the clinical and laboratory teams who collected and analyzed the samples. We are grateful to those who collaborated on the original clinical study in which the samples analyzed herein were collected. We are also grateful to R. Autrey, B. Johnson and K. Berquist for their administrative expertise at the University of Florida. We also thank Dr. James Platts-Mills for his guidance on computing attributable fractions. EJN was the PI and obtained IRB approval at the University of Florida. Associated protocol numbers/registrations are: University of Florida IRB 201703246 and the Comité National de Bioéthique (National Bioethics Committee of Haïti) Ref:1718-35. This collective research infrastructure and support was invaluable to the success of this study.

## Funding

This work was supported by the National Institutes of Health [DP5OD019893] to EJN and [R01 AI135115] to DTL as well as the Children’s Miracle Network. The funders had no role in study design, data collection and analysis, decision to publish, or preparation of the manuscript.

Conceptualization: MAS, DTL, ATM, CB, MBK, EJN

Methodology: IS, SRK, EC, YC, LB, MAS, DTL, SM, ATM, MBK, EJN

Investigation: IS, SRK, YC, EC, LB, CB, MBK, EJN

Visualization: IS, SRK, MK, EJN

Funding acquisition: DTL, EJN

Project administration: YC, MBK, EJN

Supervision: YC, MAS, VBR, CB, MBK, EJN

Formal analysis: IS, MBK, EJN

Resources: MAS, VBR, DTL, ATM, EJN

Data Curation: IS, SRK, YC, EC, LB, MBK

Writing – original draft: IS, SRK, MBK, EJN

Writing – review & editing: YC, EC, LB, MAS, SM, VBR, DTL, ATM, CB, EJN

## Competing interests

Authors declare that they have no competing interests.

## Data and materials availability

All data produced in the present study are available upon reasonable request to the authors.

## Code availability

The computer code to reproduce data analyses is available upon request.

## Notes

### Competing Interest Statement

The authors have declared no competing interest.

### Author Declarations

IRB of University of Florida gave ethical approval for this work (IRB201703246). IRB of Comité National de Bioéthique (National Bioethics Committee of Haïti) gave ethical approval for this work (Ref:1718-35).

### Summary of Updates

This version of the manuscript has been revised to update the title page.

