## Supplemental Figures for "Diarrheal disease attributed to *Shigella* spp. and Enteroinvasive *Escherichia coli* among children at households in rural Haïti: A case-control study"

### SUPPLEMENTARY MATERIALS

**Figures:** S1-S4

### SUPPLEMENTAL FIGURES

### FIGURE S1

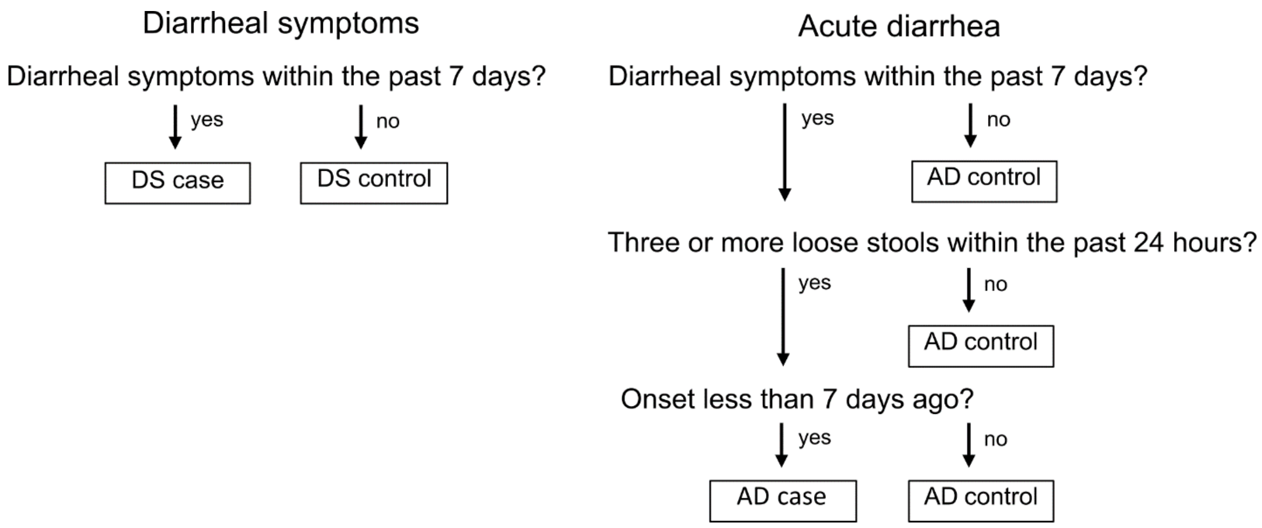


**Fig S1. Case definitions for diarrheal symptoms (DS) and acute diarrhea (AD).**

**FIGURE S2**

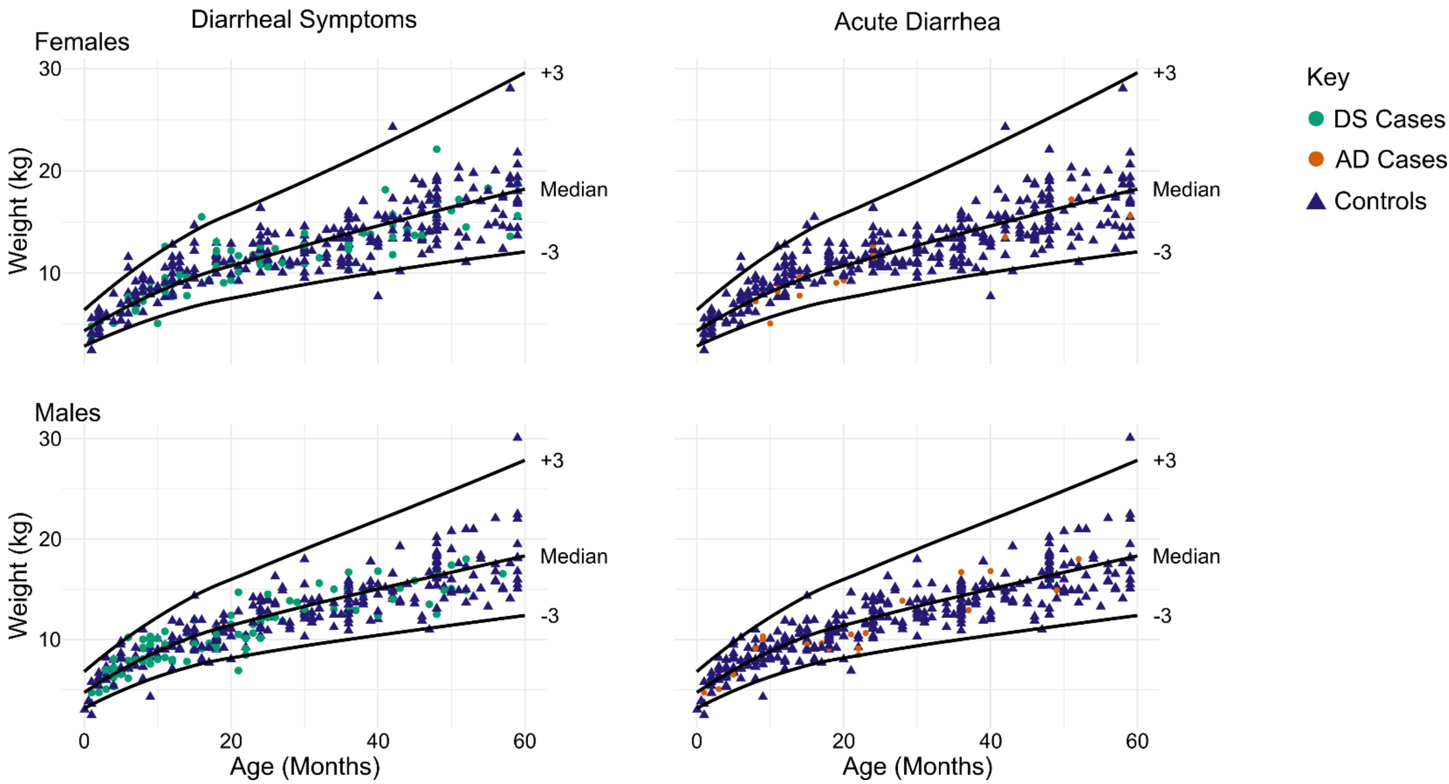


**Fig S2. Weight-for-age measurements.** Sex- specific weight-for-age and Z-scores are displayed for all participants stratified by both the DS and AD case definitions. Z-scores ≤ -3 are considered severely underweight. DS=Diarrheal symptoms; answered ‘YES’ to “diarrheal symptoms within the last 7 days”. AD=Acute diarrhea; answered ‘YES’ to “diarrheal symptoms within the last 7 days”, and “three or more loose stools in the past 24 hours”, and “onset less than 7 days ago”.

### FIGURE S3

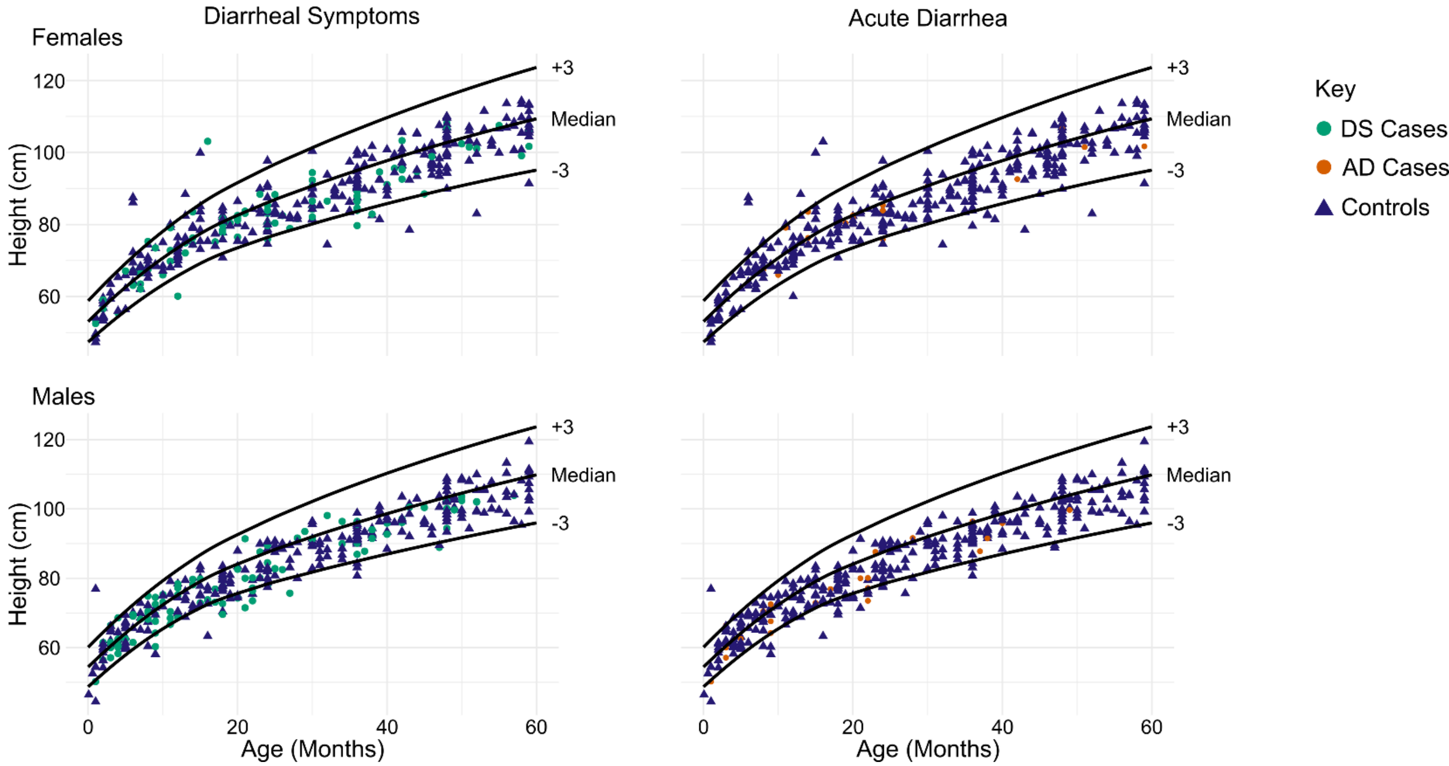


**Fig S3. Height-for-age measurements.** Sex- specific height-for-age and Z-scores are displayed for all participants stratified by both the DS and AD case definitions. DS=Diarrheal symptoms; answered 'YES' to "diarrheal symptoms within the last 7 days". AD=Acute diarrhea; answered 'YES' to "diarrheal symptoms within the last 7 days", and "three or more loose stools in the past 24 hours", and "onset less than 7 days ago".

**FIGURE S4**

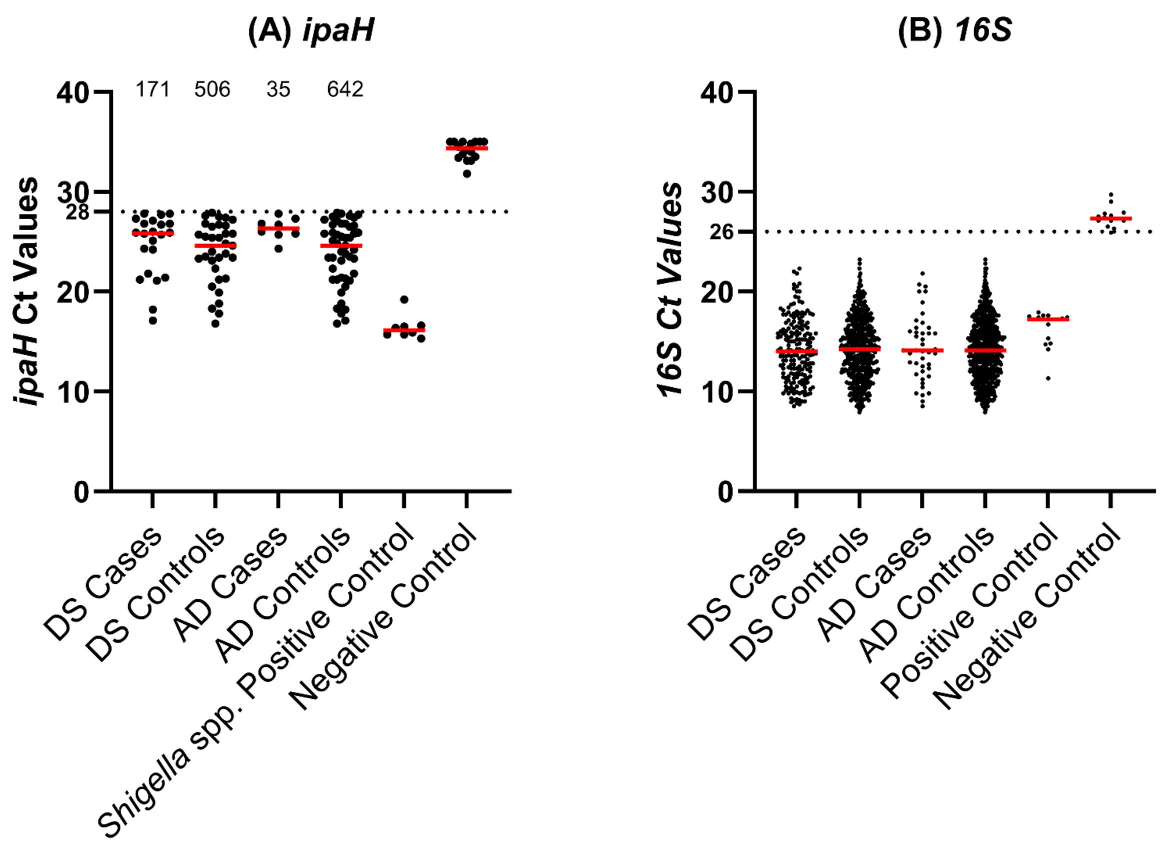


**Fig S4. *ipaH* and *16S* Ct value distributions for positive samples.** (A) The distribution of positive Ct values for *ipaH* (Ct <28) and *16S* (<26). Median Ct values for *ipaH* were 25.9 24.6, 26.35 24.6, 16.15, and 34.35 for DS Cases, DS Controls, AD Cases, and AD controls, *Shigella* spp. positive control, and negative control, respectively. Negative samples for DS cases, DS controls, AD cases, and AD controls for the *ipaH* primer are enumerated at the top of the figure. (B) Median Ct values for *16S* were 14, 14.2, 14.1, 14.1, 17.2, and 27.3, respectively. DS=Diarrheal symptoms; answered 'YES' to "diarrheal symptoms within the last 7 days". AD=Acute diarrhea; answered 'YES' to "diarrheal symptoms within the last 7 days", and "three or more loose stools in the past 24 hours", and "onset less than 7 days ago".
